# Ruxolitinib As a Salvage Therapy for Acute and Chronic Graft-versus-Host Disease in Children and Young Adults: a single institution experience

**DOI:** 10.1101/2021.10.29.21265670

**Authors:** Melissa Mavers, Edna Klinger, David C Shyr, Ami J Shah, Alice Bertaina, Sandeep Soni

**Affiliations:** Division of Hematology, Oncology, Stem Cell Transplantation and Regenerative Medicine, Department of Pediatrics, Stanford University School of Medicine, Stanford CA; Bass Center for Childhood Cancer and Blood Diseases, Stanford Children’s Health, Palo Alto, CA; Department of Pediatrics, Division of Bone Marrow Transplant, University of California San Francisco, San Francisco, CA

**Keywords:** graft-versus-host disease, acute graft-versus-host disease, chronic graft-versus-host disease, steroid-refractory, ruxolitinib, hematopoietic stem cell transplantation

## Abstract

**Background:** Ruxolitinib, a Janus kinase (JAK)-1/JAK-2 inhibitor that reduces T-cell activation and proliferation leading to a potent anti-inflammatory effect, was recently approved to treat steroid-refractory acute graft-versus-host disease (aGvHD) and is under study for chronic GvHD (cGvHD). However, there are few reports on its use in pediatric and young adult patients with GvHD.

**Methods:** We retrospectively report our single center experience with ruxolitinib in 15 patients aged 11-29 years: 5 patients with steroid refractory/steroid dependent aGvHD and 10 patients with cGvHD who had failed at least one systemic therapy.

**Results:** In the aGvHD group, ruxolitinib led to an overall response rate (ORR) of 40% at day 28 and 80% with longer follow-up (complete response, CR 60%), with a median of 21 days (5-74) to achieve a response. In the cGvHD group (9 evaluable patients), we observed an ORR of 67% and a CR of 22%, with a median of 78 days (35-180) to achieve a response. Adverse events included cytopenias, gastrointestinal symptoms, infections, and an allergic reaction attributable to ruxolitinib.

**Conclusions:** Overall, our results show that ruxolitinib is an effective salvage therapy for severe GvHD in pediatric and young adult patients. The toxicities noted warrant adequate antimicrobial prophylaxis and close monitoring of blood counts.

## INTRODUCTION

Graft-versus-host disease (GvHD) remains a leading cause of morbidity and mortality following allogeneic hematopoietic cell transplantation (HCT)^1,2^, and represents a major limitation to successful outcomes, affecting the risk-benefit profile of HCT, especially its application to non-malignant diseases. Both acute (a-) and chronic (c-) GvHD remain difficult to treat, particularly if unresponsive to steroids, to date the gold standard of GvHD treatment.

Recently ruxolitinib was shown to be efficacious in treating steroid-refractory (SR) aGvHD^3–5^, and subsequently became the first drug to receive FDA approval for this indication. Ruxolitinib functions through selective inhibition of Janus kinase (JAK) 1/2, which leads to a reduction in signaling through the common gamma chain and suppression of T-cell proliferation, activation, and survival^6^. Suppression of aGvHD by ruxolitinib was first shown in a mouse model, highlighting the abrogation of interferon gamma receptor signaling in T cells leading to reduced CXCR3 expression and decreased trafficking to GvHD target organs^7^. Regulatory effects of ruxolitinib have also been reported on innate immune cell populations, such as dendritic cells and natural killer cells^8,9^. Collectively, these suppressive effects interrupt the cycle of immune cell recruitment to the damaged tissues post-transplant and their proliferation and activation which would otherwise cause further tissue damage, the hallmark of aGvHD^10–12^. Ruxolitinib is promising for the treatment of cGvHD as well^3,13,14^. Recently, preliminary results from a randomized phase III trial demonstrated superior efficacy of ruxolitinib versus best available therapy in SR cGvHD (REACH3)^15^. This study included only 12 pediatric patients, and it is not clear how many were on the ruxolitinib arm, as the full results have not been published to date. This, combined with the limited data currently available regarding the use of ruxolitinib in the pediatric and YA population^16–20^, highlights the need for ongoing reporting of results with ruxolitinib use in this age group. Herein, we report our early experience of using ruxolitinib in pediatric and YA patients with aGvHD and cGvHD at a single center, the first such report to include patients with cGvHD treated at a pediatric center in the United States.

## PATIENTS AND METHODS

### Patients and Approval

The charts of all patients who received ruxolitinib for GvHD following allogeneic HCT at Lucile Packard Children’s Hospital between 8/1/2018 and 8/31/2019 were analyzed retrospectively after obtaining Institutional Review Board approval (IRB-53278). Patients were followed through the data cut-off of 1/31/2020, allowing a follow-up period of 6-18 months to gauge response and collect data on adverse events.

### Definitions and Grading of GvHD

Acute GVHD was diagnosed and graded according to the Modified Glucksberg scale^21^ as per institutional practice. Whenever possible, the diagnosis was corroborated with tissue biopsies. SR was defined as progression of symptoms after 3-5 days of standard steroid treatment (2mg/kg per day of IV methylprednisolone or equivalent prednisone) or lack of response after 5-7 days of continued steroid use. Steroid dependent (SD) was defined as a flare of symptoms during the period of steroid wean. Chronic GvHD was scored as limited or extensive based on Seattle criteria^22^.

### Treatment with Ruxolitinib

Patients with SR/SD aGvHD who were in good performance status with ability to take oral medications and patients with cGvHD who were non-responders to at least one systemic therapy were started on ruxolitinib. The agents targeting GvHD which each patient had received prior to starting ruxolitinib were recorded, including GVHD prophylaxis (if continued or restarted for GVHD treatment), excluding topical medications. Ongoing immune suppressive medications were continued or discontinued at the discretion of the stem cell transplant team at the time of starting ruxolitinib. Since there is no recommended dose for ruxolitinib in pediatric patients, treatment was initiated with 5mg twice daily (BID) and titrated per patient tolerance and clinical response as an institutional plan. The dose was not increased beyond a maximum of 20mg BID, per standard adult dosing practices. Patients who responded adequately were weaned off ruxolitinib over 2-4 weeks prior to discontinuation. CBC and labs to monitor for liver, renal and metabolic parameters were routinely performed.

### Infection Prophylaxis

All patients with acute GVHD received antiviral, antifungal, and Pneumocystis jiroveci pneumonia (PJP) prophylaxis as per institutional practice. Patients with chronic GvHD were on variable antiviral, antifungal, and PJP prophylaxis depending on the transplant indication, time post-transplant, lymphocyte mitogen responses, and viral reactivation status.

### Response

For aGvHD, a partial response (PR) was defined as a reduction of at least one stage in GvHD severity in any organ, without worsening of any other organ and a complete response (CR) was defined as the absence of all signs or symptoms of GvHD. For patients with gastrointestinal (GI) aGvHD upgraded in severity due to abdominal pain, a PR was defined as a reduction in pain medication requirements. Overall response rate (ORR) was calculated as the sum of PR and CR. Day 28 response and best response at any time in the follow-up period were analyzed. Patients were determined to have no response (NR) at day 28 if they had stable disease or treatment failure (TF) if they had progressive disease at any time.

For cGvHD, a PR was defined as reduction in concomitant immune suppressive drugs (or the frequency of extracorporeal photopheresis procedures) by at least 50% at any time, along with a significant improvement in cGvHD manifestations. A CR was defined as the resolution of signs or symptoms of cGvHD without requiring additional immune suppression during follow-up. Patient was considered a TF if additional immune suppression was added at any time during the follow-up period.

Time to initial response with ruxolitinib was also analyzed for both of the cohorts.

### Toxicities and infections

All patient charts were analyzed for side effects, toxicities, incidence of major infections, and laboratory parameters during the period of treatment with ruxolitinib. Dose-limiting toxicities as well as infections or viral reactivations requiring treatment were captured.

## RESULTS

### Patient Characteristics

A total of 15 patients aged 11-29 years old received ruxolitinib for SR/SD aGvHD (n=5) or for cGvHD (n=10) (Table 1). Of note, one patient was diagnosed with late onset aGvHD (on day +167) and was evaluated in the aGvHD group. Patient demographics and transplant characteristics are shown in Table 1. Of note, four patients had received a second transplant. Majority of patients had received initial GvHD prophylaxis with a calcineurin inhibitor and methotrexate or mycophenolate mofetil, while two patients had received sirolimus and mycophenolate mofetil, and one patient who received a αβ T/CD19^+^ B-cell depleted graft had received no prophylaxis (Table 1). Patients were followed for a median of 310 days (range 164-691) after starting ruxolitinib.

**Table 1.**
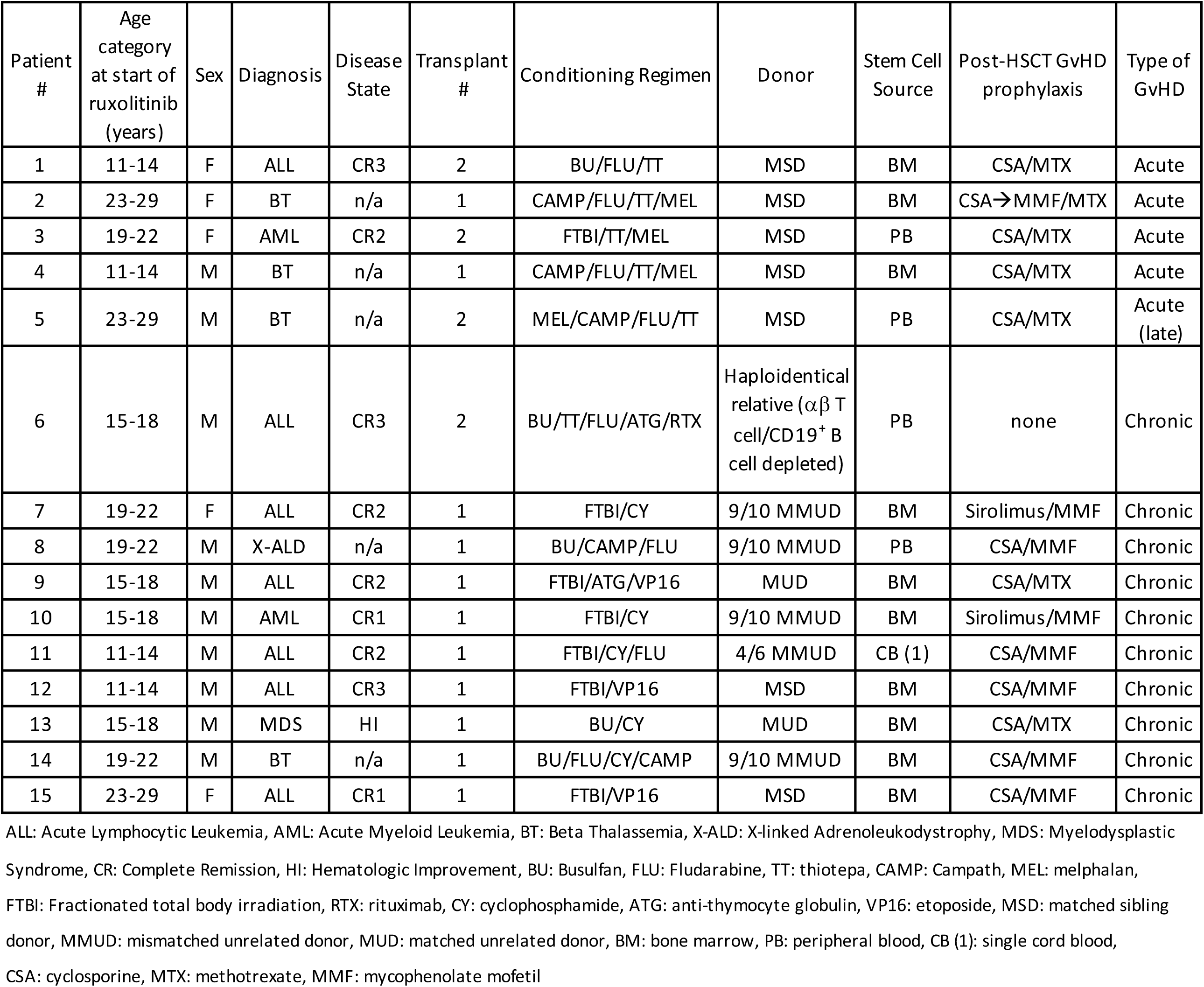
Patient Demographics

| Patient # | Age category at start of ruxolitinib (years) | Sex | Diagnosis | Disease State | Transplant # | Conditioning Regimen | Donor | Stem Cell Source | Post-HSCT GvHD prophylaxis | Type of GvHD |
| --- | --- | --- | --- | --- | --- | --- | --- | --- | --- | --- |
| 1 | 11-14 | F | ALL | CR3 | 2 | BU/FLU/TT | MSD | BM | CSA/MTX | Acute |
| 2 | 23-29 | F | BT | n/a | 1 | CAMP/FLU/TT/MEL | MSD | BM | CSA→MMF/MTX | Acute |
| 3 | 19-22 | F | AML | CR2 | 2 | FTBI/TT/MEL | MSD | PB | CSA/MTX | Acute |
| 4 | 11-14 | M | BT | n/a | 1 | CAMP/FLU/TT/MEL | MSD | BM | CSA/MTX | Acute |
| 5 | 23-29 | M | BT | n/a | 2 | MEL/CAMP/FLU/TT | MSD | PB | CSA/MTX | Acute (late) |
| 6 | 15-18 | M | ALL | CR3 | 2 | BU/TT/FLU/ATG/RTX | Haploidentical relative ($\alpha\beta$ T cell/CD19<sup>+</sup> B cell depleted) | PB | none | Chronic |
| 7 | 19-22 | F | ALL | CR2 | 1 | FTBI/CY | 9/10 MMUD | BM | Sirolimus/MMF | Chronic |
| 8 | 19-22 | M | X-ALD | n/a | 1 | BU/CAMP/FLU | 9/10 MMUD | PB | CSA/MMF | Chronic |
| 9 | 15-18 | M | ALL | CR2 | 1 | FTBI/ATG/VP16 | MUD | BM | CSA/MTX | Chronic |
| 10 | 15-18 | M | AML | CR1 | 1 | FTBI/CY | 9/10 MMUD | BM | Sirolimus/MMF | Chronic |
| 11 | 11-14 | M | ALL | CR2 | 1 | FTBI/CY/FLU | 4/6 MMUD | CB (1) | CSA/MMF | Chronic |
| 12 | 11-14 | M | ALL | CR3 | 1 | FTBI/VP16 | MSD | BM | CSA/MMF | Chronic |
| 13 | 15-18 | M | MDS | HI | 1 | BU/CY | MUD | BM | CSA/MTX | Chronic |
| 14 | 19-22 | M | BT | n/a | 1 | BU/FLU/CY/CAMP | 9/10 MMUD | BM | CSA/MMF | Chronic |
| 15 | 23-29 | F | ALL | CR1 | 1 | FTBI/VP16 | MSD | BM | CSA/MMF | Chronic |

ALL: Acute Lymphocytic Leukemia, AML: Acute Myeloid Leukemia, BT: Beta Thalassemia, X-ALD: X-linked Adrenoleukodystrophy, MDS: Myelodysplastic Syndrome, CR: Complete Remission, HI: Hematologic Improvement, BU: Busulfan, FLU: Fludarabine, TT: thiotepa, CAMP: Campath, MEL: melphalan, FTBI: Fractionated total body irradiation, RTX: rituximab, CY: cyclophosphamide, ATG: anti-thymocyte globulin, VP16: etoposide, MSD: matched sibling donor, MMUD: mismatched unrelated donor, MUD: matched unrelated donor, BM: bone marrow, PB: peripheral blood, CB (1): single cord blood, CSA: cyclosporine, MTX: methotrexate, MMF: mycophenolate mofetil

### Acute GvHD Characteristics and Response

Four of the 5 patients with aGvHD had severe SR disease (Grade III) and one (#1) had SD disease (flare of skin rash and rising bilirubin during steroid taper) as the indication for starting ruxolitinib. Patients had received 2-5 agents targeting GvHD prior to the addition of ruxolitinib, which was initiated after a median of 21 days (range 6-94) of systemic steroid use and after a median of 23 days (range 7-102) from aGvHD diagnosis. All patients tolerated oral ruxolitinib well, including those with GI aGvHD.

After 28 days of ruxolitinib treatment, PR was observed in 2 patients (40%) with no CR, while an ORR of 80%, with a CR of 60% (N=3) was observed with ruxolitinib after longer treatment follow-up (Table 2). Patients took a median of 21 days (range: 5-74) to achieve initial response and remained on ruxolitinib for a median of 62 days (range: 27-332). Median time to CR (N=3) was 54 days (range: 41-226). One patient (#4) who had achieved CR discontinued ruxolitinib, and another who achieved CR (#1) was still on ruxolitinib with cyclosporin being tapered at the time of data cut-off (day +220 following transplant). Two patient deaths occurred in the acute GvHD group: one (#5) had achieved a CR and subsequently had a severe flare of GI GvHD (while still on ruxolitinib) and died following intestinal perforation and multisystem organ failure, and the other (#3) had progressive aGvHD and was deemed a treatment failure. Two responders (one PR and one CR) subsequently developed cGvHD. Both of these patients were restarted on ruxolitinib for cGvHD.

**TABLE 2.**
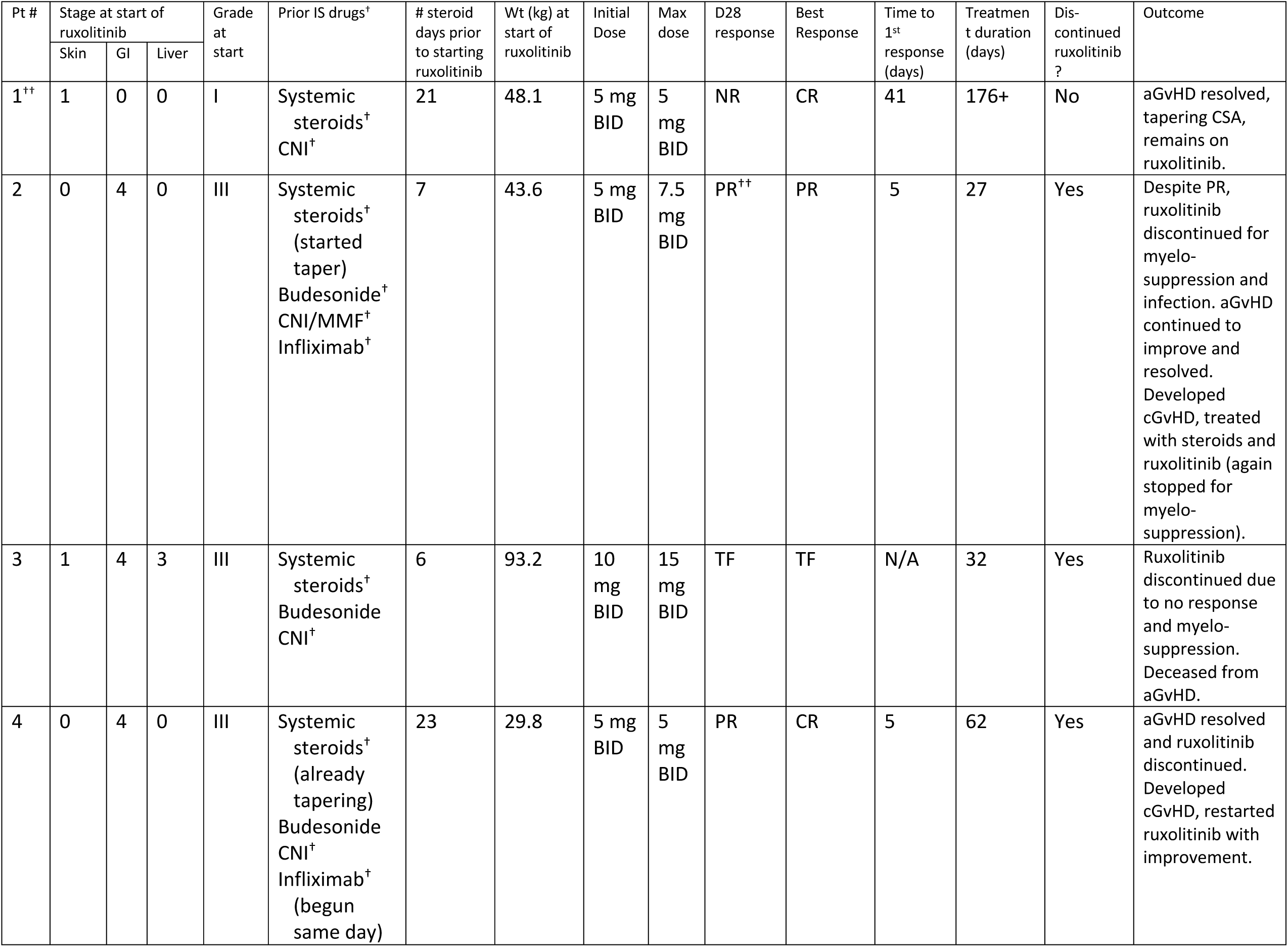
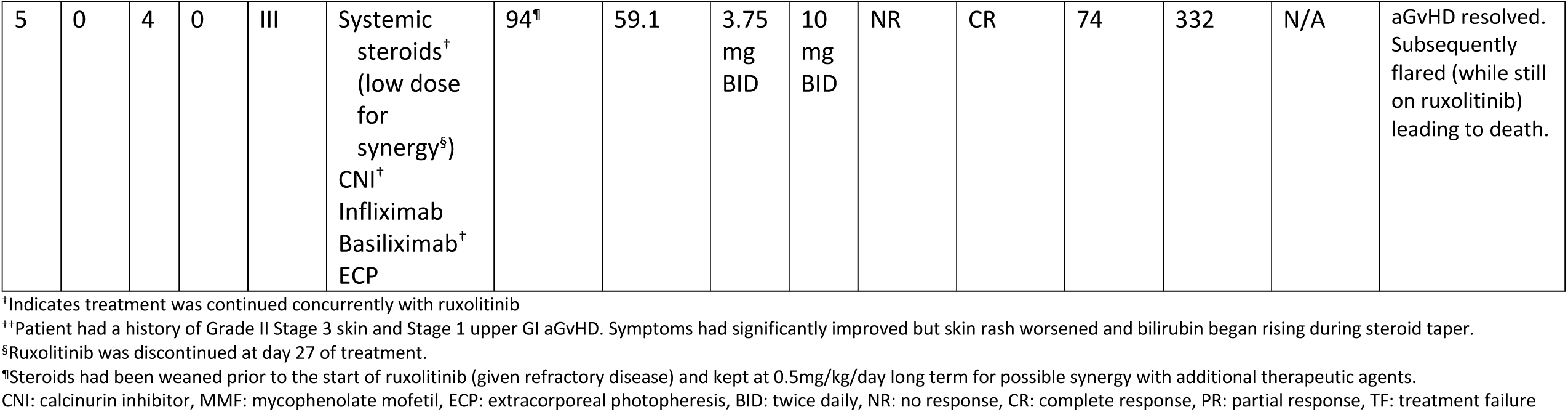
Acute GvHD and Ruxolitinib Use

### Chronic GvHD Characteristics and Response

Ruxolitinib was administered for cGvHD in 10 patients—3 of whom had limited disease and 7 with extensive disease (Table 3). All patients had previously received 1-6 immunosuppressive agents, with 4 patients remaining on prolonged systemic steroids for control of cGvHD at the time ruxolitinib was started. Of note, in one of our patients (#12), all other prior immune suppression was discontinued at the time of starting ruxolitinib and response was evaluated based on clinical improvement in cGvHD manifestations. One patient had ruxolitinib discontinued after 7 days due to an allergic reaction (see below) and therefore, was not considered evaluable for response.

**TABLE 3.** Chronic GvHD and Ruxolitinib Use

| Pt # | Severity at start of ruxolitinib | Organs Involved at start of ruxolitinib | Prior IS drugs for cGvHD<sup>+</sup> | # steroid days prior to starting ruxolitinib | Wt (kg) at start of ruxolitinib | Initial Dose | Max dose | Response | Time to 1<sup>st</sup> response (days) | Treatment duration (days) | Dis-continued ruxolitinib? | Outcome |
| --- | --- | --- | --- | --- | --- | --- | --- | --- | --- | --- | --- | --- |
| 6 | Extensive | skin, polyserositis | Systemic steroids<sup>+</sup> (started taper)<br>Budesonide<sup>+</sup> (decreased dose)<br>Methotrexate | 21 | 51.4 | 5 mg BID | 5 mg BID | TF | NA | 259 | Yes | Tapered off steroids, cGvHD later flared (on ruxolitinib), steroids restarted, flare resolved, steroids tapered off, and ruxolitinib discontinued for resolution of cGvHD. |
| 7 | Extensive | skin, mouth | Budesonide<sup>+</sup><br>CNI<sup>+</sup><br>Infliximab | NA | 65.4 | 5 mg BID | 10 mg qAM, 5 mg qHS | PR | 105 | 310+ | No | cGvHD continues to gradually improve |
| 8 | Limited | skin | Systemic steroids<sup>++</sup><br>CNI<sup>+</sup> (already tapering) | NA<sup>++</sup> | 65.1 | 5 mg BID | 5 mg BID | CR | 72 | 164+ | No | cGvHD resolved, CSA tapered off, plan ruxolitinib taper |
| 9 | Extensive | skin, upper GI | CNI<sup>+</sup> (started taper) | NA | 41.1 | 5 mg daily | 5 mg BID | CR | 28 | 196 | Yes | cGvHD resolved and ruxolitinib discontinued. |
| 10 | Extensive | skin, mouth | Systemic steroids<sup>+</sup> (started taper)<br>CNI<sup>+</sup> | 35 | 46.3 | 5 mg BID | 10 mg BID | PR | 84 | 436+ | No | cGvHD nearly resolved, steroids tapered off, plan ruxolitinib taper |
| 11 | Extensive | skin | Systemic steroids<br>CNI<br>ECP<br>Sirolimus | NA (stopped >1 year prior) | 28.5 | 5 mg BID | 10 mg BID | TF | NA | 264+ | No | cGvHD progressed to include GI, increased ruxolitinib, added steroids |
| 12 | Extensive | skin | ECP<br>Imatinib | NA | 71.3 | 5 mg<br>BID | 10<br>mg<br>BID | PR<sup>§</sup> | 35<sup>§</sup> | 230 | Yes | Primary disease<br>relapsed; ruxolitinib<br>discontinued. |
| 13 | Extensive | skin, lungs | Systemic<br>steroids<br>ECP<sup>†</sup><br>Imatinib<br>Ibrutinib<br>Azathioprine<br>Perfenidone<sup>†</sup> | NA<br>(stopped<br>>1<br>year<br>prior) | 61 | 2.5<br>mg<br>BID | 15<br>mg<br>qAM<br>, 10<br>mg<br>qHS | PR | 180 | 525+ | No | cGvHD continues to<br>gradually improve |
| 14 | Limited | skin | Systemic<br>steroids<sup>†</sup><br>CNI<br>MMF<br>ECP<br>Sirolimus | 1655 | 59 | 10<br>mg<br>BID | 20<br>mg<br>BID | TF | NA | 487+ | No | cGvHD flared<br>requiring additional<br>immune suppression,<br>steroids<br>subsequently<br>tapered off, now<br>tapering other<br>immune suppression |
| 15 | Limited<sup>¶</sup> | mouth<sup>¶</sup> | Systemic<br>steroids<sup>†</sup><br>CNI<br>MMF | 223 | 46.3 | 5 mg<br>BID | 5 mg<br>BID | NE | NA | 7 | Yes | Ruxolitinib<br>discontinued for<br>severe allergic<br>reaction. Ongoing<br>cGvHD. |

<sup>†</sup>Indicates treatment was continued concurrently with ruxolitinib
<sup>††</sup>Patient on long-term low dose prednisone for adrenal insufficiency pre- and post-transplant
<sup>§</sup>Response based on improvement in symptoms as all other immune suppression stopped when ruxolitinib started.
<sup>¶</sup>Patient also with remote history of extensive chronic GvHD of skin, joints, and eyes which had resolved and was off all IS for over 2 years.
CNI: calcinurin inhibitor, MMF: mycophenolate mofetil, ECP: extracorporeal photopheresis, BID: twice daily, CR: complete response, PR: partial response, TF: treatment failure, NE: Not evaluable

Of the 9 patients evaluable for response, an ORR of 67%, with a CR in 2 (22%) was observed. Initial response was observed after a median of 78 days (range: 28-180) and patients remained on ruxolitinib treatment for a median of 264 days (range 164-525). Two patients discontinued ruxolitinib due to resolution of cGvHD (#6 and 9) and one discontinued ruxolitinib due relapse of the underlying malignancy (#12). Of the 3 evaluable patients receiving systemic steroids for control of cGvHD at the time ruxolitinib was started (#6, 10, and 14), all were able to taper off steroids, despite two being considered ruxolitinib treatment failures as described below. Three patients required the use of additional immune suppressive medication and were considered treatment failures. In one of these patients (#6), ruxolitinib use led to near resolution of cGvHD and steroids were discontinued, but the skin GvHD flared while still on ruxolitinib and steroids were restarted. The steroids were subsequently weaned off with no further flares, and ruxolitinib was then discontinued due to resolution of cGvHD. Another patient (#14) similarly had a flare of GvHD requiring additional immune suppression which was being tapered at the time of data cut-off. The third treatment failure patient (#11) had progressive GvHD, with involvement of an additional site of disease, leading to an increase in ruxolitinib dose and addition of steroids. No mortality was observed in the cGVHD cohort through the time of data cut-off.

### Toxicities and Infections

One severe adverse event in the cGvHD group was observed in a patient (#15) who developed Stevens-Johnson syndrome requiring hospitalization, 7 days after starting ruxolitinib. This was attributed to ruxolitinib due to lack of exposure to any other new medications. Ruxolitinib was discontinued and the patient rapidly improved with medical management.

Of the remaining 14 patients who tolerated ruxolitinib, 6 (43%) were noted to have dose-limiting cytopenias, at a median dose of 5mg BID (range 5mg BID to 15mg BID, Table 4). Four of these patients were in the aGvHD cohort and 2 in the cGvHD cohort, although only 2 patients required discontinuation of ruxolitinib due to myelosuppression. Several of these patients developed cytopenias as their dose was being escalated, which improved after dose reduction. Additionally, two patients (one each in the aGvHD and cGvHD cohorts) required a dose reduction due to gastrointestinal symptoms, including significant increase in abdominal pain, increased gas, and increased frequency (but reduced volume) of stools. Of note, both patients also had developed cytopenias requiring dose reduction. GI symptoms also resolved with the reduction in dose. No other non-infectious adverse events were noted, including no dose-limiting increase in liver enzymes or triglycerides.

**TABLE 4.** Dose-Limiting Toxicities

| GvHD Group | Evaluable Patients (N) | Dose Limiting Toxicity | Patients (N) | Patients (%) |
| --- | --- | --- | --- | --- |
| Acute | 5 | Cytopenias | 4 | 80% |
|  |  | GI toxicity | 1 | 20% |
| Chronic | 9 | Cytopenias | 2 | 22% |
|  |  | GI toxicity | 1 | 11% |

Four patients (80%) in the aGvHD cohort and 4 (44%) in the cGvHD cohort, developed at least one episode of infection during treatment with ruxolitinib (Table 5), though a majority of the patients were on additional immune suppression as well (Tables 2 and 3). Viral infections and reactivations were particularly predominant in both the aGvHD and cGvHD cohorts (Table 5). Additionally, one patient in our aGvHD cohort (#3) developed a probable fungal pulmonary infection (based on chest CT findings and rising galactomannan level), though this was diagnosed within 2 days of starting ruxolitinib.

**TABLE 5.** Infections During Ruxolitinib Treatment

| Patient # | Acute or Chronic GvHD | Infections/reactivations |
| --- | --- | --- |
| 1 | Acute | none |
| 2 | Acute | Adenoviral infection (cystitis, enteritis)<br>CMV reactivation<br>Enterobacter cloacae and Staphylococcus aureus abscess |
| 3 | Acute | BK+ hematuria (improved without treatment)<br>Probable fungal pulmonary infection (diagnosed within 2 days of starting ruxolitinib) |
| 4 | Acute | MSSA bacteremia |
| 5 | Acute (late) | Gastroenteritis (likely viral)<br>Klebsiella pneumonia osteomyelitis and sepsis<br>MSSA and Enterobacter sepsis |
| 6 | Chronic | none |
| 7 | Chronic | Respiratory Syncytial Virus upper respiratory infection<br>Rhinovirus/Enterovirus upper respiratory infection |
| 8 | Chronic | none |
| 9 | Chronic | none |
| 10 | Chronic | Respiratory Syncytial Virus and Influenza A infection<br>Rhinovirus/Enterovirus upper respiratory infection<br>Bilateral acute otitis media (presumed bacterial) |
| 11 | Chronic | none |
| 12 | Chronic | Unspecified upper respiratory infection |
| 13 | Chronic | Unspecified upper respiratory infection (x2)<br>Norovirus gastroenteritis<br>Escherichia coli bacteremia |
| 14 | Chronic | none |
| 15 | Chronic | none |

CMV: Cytomegalovirus, EBV: Epstein Barr virus, MSSA: Methicillin-sensitive Staphylococcus aureus

Bacterial infections were noted in 60% of patients with aGvHD (including 2 episodes in one patient) and in 20% of patients with cGvHD. There was no clear association between use of antibacterial prophylaxis and absence of bacterial infection, as 2 of the 3 patients with aGvHD who developed bacterial infections were on prophylaxis with levofloxacin and both patients with cGvHD were on daily sulfamethoxazole/trimethoprim.

## DISCUSSION

In this report, we sought to add our experience to the relatively few descriptions of ruxolitinib use in pediatric and young adult patients with GvHD, including the first report using ruxolitinib in patients with cGvHD treated at a pediatric center in the United States. All patients tolerated taking ruxolitinib by mouth, including those with GI aGvHD. The day 28 response rate in our patients with SR/SD-aGvHD (40% ORR, 0% CR) was lower than that reported in the Phase 3 REACH2 trial in which 149 adult patients and 5 pediatric patients had a 62% ORR (34% CR) at day 28^5^. However, our results were similar to the week 4 outcomes published by Khandelwal et al. on 13 pediatric patients with SR-aGvHD (45% ORR, 9% CR)^16^. The best response achieved in our cohort of patients with SR/SD-aGvHD (80% ORR) was similar to that reported in other pediatric and adult studies^3,4,16–20^. Though comparisons are difficult given the small number of patients with aGvHD in our study, the lower response rate at day 28 in pediatric patients as compared to adults may be due to the uncertainty of dosing in pediatric patients who may have increased clearance of the drug, and as we gradually increased the dose of ruxolitinib over time, this practice may take longer to achieve therapeutic levels compared to adult patients. Indeed, patients with SR/SD-aGvHD required a median of 21 days (range: 5-74) to achieve initial response and 54 days (range: 41-226) to achieve CR. This suggests that continued and sustained treatment with ruxolitinib is required to assess adequate response, and discontinuation (designating treatment failure) should not be done prematurely. Importantly, we saw a response even in our patients with SR GI aGvHD (75% ORR) suggesting ruxolitinib is absorbed systemically in these patients.

In our cGvHD group, our CR (22%) was similar to that reported in other pediatric studies, while our ORR (67%) was lower^17,18,20^. The reduction in ORR was not due to increased disease severity, as this was similar to other studies, but may reflect the small number of heterogenous patients in these reports, differences in co-administration of other immunosuppressive drugs, and/or variations in response criteria. Because of the retrospective nature of this study, we were unable to re-score patients using NIH consensus criteria. Importantly, all evaluable patients with cGVHD in our study who were on steroids at the time of starting Ruxolitinib were able to wean off steroids, highlighting the steroid sparing effect of ruxolitinib in this population. This is an important consideration, as prolonged use of steroids is related to multiple complications in the pediatric population, especially related to bone health and growth.

It is also important to note the time to response for patients in the cGvHD cohort. In these patients, initial response was observed after a median of 78 days (range: 28-180). This again suggests that it may take a prolonged period of time to see the therapeutic effect of ruxolitinib and that early lack of response in patients with cGvHD is not an indication to discontinue the drug as long as it remains well tolerated. Some patients who had an initial response with ruxolitinib and were weaned off steroids had flare of symptoms during follow-up. Further studies with additional biomarkers are needed in the pediatric population to delineate a better weaning plan for immunosuppressive agents in conjunction with ruxolitinib.

The majority of toxicities we report are similar to those published in earlier studies, including cytopenias and potentially increased risk of infection^3–5^. Doses need to be titrated based on blood cell counts. Interestingly, we did not observe any liver toxicity in our patients, including no ruxolitinib-related increase in triglycerides^23^. Additionally, we report a significant allergic reaction attributable to ruxolitinib in a GvHD patient.

Although this report is limited by its retrospective nature, lack of NIH consensus grading for cGvHD, and small number of patients, our results suggest that ruxolitinib is a good candidate as a first line agent for SR/SD-aGvHD and as a steroid sparing agent in cGvHD for pediatric and young adult patients. Many patients responded despite having failed multiple other therapies, suggesting that earlier use of ruxolitinib may lead to enhanced responses and avoidance or earlier tapering of other immunosuppressive therapies. Importantly, our study also corroborates the increased risk of infections in patients with GvHD and on ruxolitinib and other immunosuppressive therapies, and the need for prophylactic medications should be carefully evaluated. In addition, more clear pharmacokinetic data in pediatric patients is needed to guide dosing and weaning plans. A single-arm international clinical trial (currently without open sites in the United States) aims to evaluate the pharmacokinetics, safety, and activity of ruxolitinib in pediatric patients with newly diagnosed and SR-aGvHD (NCT03491215) and will represent an important step forward in further understanding the use of ruxolitinib in this population.

## Data Availability

All data produced in the present study are available upon reasonable request to the authors, pending appropriate approval for access to clinical data.

## ACKNOWLEDGEMENTS

This work was supported by grants from the NIH (K08 HL151809 to MM) and St. Baldrick’s Foundation (Scholar Award to MM).

## Abbreviations

JAK: Janus Kinase
aGvHD: Acute Graft-versus-Host Disease
cGvHD: Chronic Graft-versus-Host Disease
ORR: Overall Response Rate
CR: Complete Response
HCT: Hematopoietic Cell Transplantation
SR: Steroid-refractory
SD: Steroid dependence
PR: Partial Response
GI: Gastrointestinal
NR: No Response
TF: Treatment Failure

